# International Multi-Specialty Expert Physician Preoperative Identification of Extranodal Extension in Oropharyngeal Cancer Patients using Computed Tomography: Prospective Blinded Human Inter-Observer Performance Evaluation

**DOI:** 10.1101/2023.02.25.23286432

**Authors:** Multidisciplinary Oropharyngeal Cancer Extra-Nodal Extension (OPC ENE) Assessment Working Group

**Affiliations:** The University of Texas MD Anderson Cancer Center, Houston, USA; University of Helsinki and Helsinki University Hospital, Helsinki, Finland; Brigham and Women’s Hospital, Dana-Farber Cancer Institute, Harvard Medical School, Boston, USA; Aalto University, Espoo, Finland; Tampere University, Faculty of Medicine and Health Technology and Tampere University Hospital, Tampere, Finland; Department of Otolaryngology Head and Neck Surgery, Baylor College of Medicine, Houston, USA

## Abstract

**Importance:** Extranodal extension (pENE) is a critical prognostic factor in oropharyngeal cancer (OPC) that drives therapeutic disposition. Determination of pENE from radiological imaging has been associated with high inter-observer variability. However, the impact of clinician specialty on human observer performance of imaging-detected extranodal extension (iENE) remains poorly understood.

**Objective:** To characterize the impact of clinician specialty on the accuracy of pre-operative iENE in human papillomavirus-positive (HPV+) OPC using computed tomography (CT) images.

**Design, Setting, and Participants:** This prospective observational human performance study analyzed pre-therapy CT images from 24 HPV+ OPC patients, with duplication of 6 scans (n=30) of which 21 were pathologically confirmed pENE. Thirty-four expert observers, including 11 radiologists, 12 surgeons, and 11 radiation oncologists, independently assessed these scans for iENE and reported human-detected radiologic criteria and observer confidence.

**Main Outcomes and Measures:** The primary outcomes included accuracy, sensitivity, specificity, area under the receiver operating characteristic curve (AUC), and Brier score for each physician, compared to ground-truth pENE. The significance of radiographic signs for prediction of pENE were determined through logistic regression analysis. Fleiss’ kappa measured interobserver agreement, and Hanley-MacNeil AUC discrimination testing.

**Results:** Median accuracy across all specialties was 0.57 (95%CI 0.39 to 0.73), with no specialty showing discriminate performance greater than random estimation (median AUC 0.64, 95%CI 0.44 to 0.83). Significant differences between radiologists and surgeons in Brier scores (0.33 vs. 0.26, p < 0.01), radiation oncologists and surgeons in sensitivity (0.48 vs. 0.69, p > 0.1), and radiation oncologists and radiologists/surgeons in specificity (0.89 vs. 0.56, p > 0.1). Indistinct capsular contour and nodal necrosis were significant predictors of correct pENE status among all specialties. Interobserver agreement was weak for all the radiographic criteria, regardless of specialty (*κ*<0.6).

**Conclusions and Relevance:** Multiobserver testing shows physician discrimination of HPV+OPC pENE on pre-operative CT remains non-different than blind guessing, with high inter-rater variability and low diagnostic accuracy, regardless of clinician specialty. While minor differences in diagnostic performance among specialties are noted, they do not significantly affect the overall poor agreement and discrimination rates observed. The findings underscore the need for further research into automated detection systems or enhanced imaging techniques to improve the accuracy and reliability of iENE assessments in clinical practice.

Visual Abstract

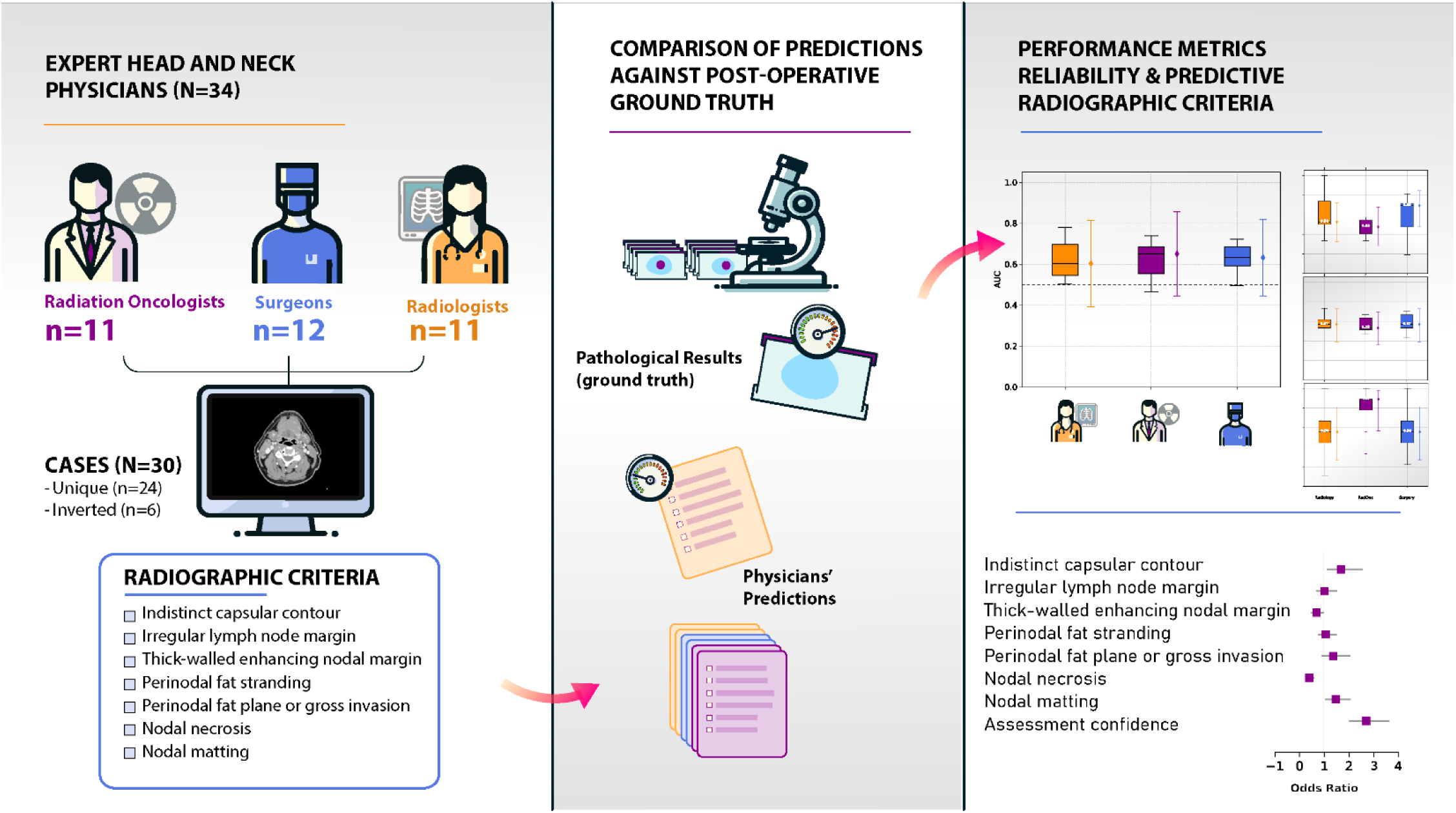

## INTRODUCTION

Extranodal extension (ENE), a phenomenon where tumor cells extend beyond the capsule of a lymph node with tumor metastasis, is among the most important adverse prognostic factors in oropharyngeal cancer (OPC), and head and neck squamous cell carcinoma (HNSCC) more broadly ^1^. ENE is often used in clinical decision-making to determine the therapeutic approach for human papillomavirus-positive (HPV+) OPC patients. While there is ambiguity regarding the impact of clinical/radiographic nodal extension in terms of chemoradiation efficacy, large-scale surgical registry data from the National Cancer Database showed that in >66,000 patients, documented pathologic ENE (pENE) was associated with an estimated 60% decrease in overall survival in patients treated surgically ^2^. The current treatment paradigms recommend adjuvant chemoradiotherapy when pENE is present ^3^. Alternatively, minimally invasive surgery, e.g., trans-oral robotic surgery, may be preferred if pENE is unlikely ^4,5^. Therefore, pre-therapy discrimination of presumptive pENE status (known as imaging-detected ENE or iENE) is crucial for appropriate treatment stratification (e.g., primary surgical or non-surgical therapy), which may have significant impacts on patient outcomes^2,6^.

The current gold-standard approach to identify nodal integrity in OPC patients involves histopathological evaluation of lymph nodes ^1^. Radiological identification of extracapsular spread using commonly available imaging modalities, such as computed tomography (CT), has long been seen as an attractive alternative for the non-invasive determination of radiographic iENE. Unfortunately, numerous studies have demonstrated that clinician-based radiological iENE as identification of pathologic extranodal extension (pENE) in OPC using radiological imaging is prone to high variability and poor discriminative performance ^7–12^. Naturally, most of these studies have specifically investigated the discriminative ability of diagnostic radiologists. However, contemporary evaluation and treatment of OPC is typically dependent on the consensus of a multidisciplinary team ^13,14^, with diverse input from clinicians specialized in radiology, surgery, and radiation oncology. Moreover, in many cases, the determination of surgery or radiotherapy (RT) as an initial treatment is driven by surgeon and radiation oncologist interpretation of imaging data in addition to radiologist assessment. Therefore, it is of vital importance to investigate and understand differences between clinical specialties in the interpretation of radiological detectability of pENE, in addition to overall human expert observer performance.

Prior work from our group has demonstrated that optimal selection of trans-oral robotic surgery with neck dissection (TORS+ND) alone vs. adjuvant radiotherapy (RT) or chemoradiation (CRT) is driven largely by the toxicity associated with adjuvant RT superimposed on surgical toxicity, which is itself driven by the probability of pENE as an indication for adjuvant CRT ^4^. Since many studies demonstrate either a substantial majority of TORS+ND cases dispositioned towards attempted non-radiotherapeutic approaches subsequently require RT or CRT ^5,15^, there appears to be a substantive optimism between pre-therapy surgical neck risk assessment and demonstrated post-operative pENE status. Put simply, quantifying cross-disciplinary physician-observer capability to effectively risk-stratify potentially operable patients based on non-invasive imaging for features to effectively identify patients for unimodality therapy is an imperative unmet need.

In this study, using a large number of clinician annotators, we prospectively benchmarked specialty-specific discriminative ability of detecting ENE/pENE in HPV+ OPC, comparing radiographic assessment on standard-of-care contrast-enhanced CT imaging to multi-pathologist-rated histopathology as the gold standard. Using various measures of discriminative performance and observer variability, we probed the underlying relationships between radiologist, surgeon, and radiation oncologist observers in their interpretation of the detectability of pENE on standard-of-care contrast CT. Additionally, we determine the relative intra- and inter-observer performance of these expert physicians through a prospective blinded *in silico* performance benchmarking assessment.

## MATERIALS & METHODS

This study followed both STrengthening the Reporting of OBservational studies in Epidemiology (STROBE) and Guidelines for Reporting Reliability and Agreement Studies (GRRAS) reporting guidelines ^16,17^. Data were collected under a HIPAA-compliant protocol approved by Institutional Review Board at The University of Texas MD Anderson Cancer Center (RCR03-0800 and PA19-0491).

### Clinician annotator/survey characteristics and data collection

Thirty-four expert clinician annotators were recruited for this prospective study: 11 radiologists, 12 surgeons, and 11 radiation oncologists. Observer characteristics are shown in **Table 1**. Strengthening the Reporting of Observational Studies in Epidemiology (STROBE) Statement guidelines.

**Table 1.**
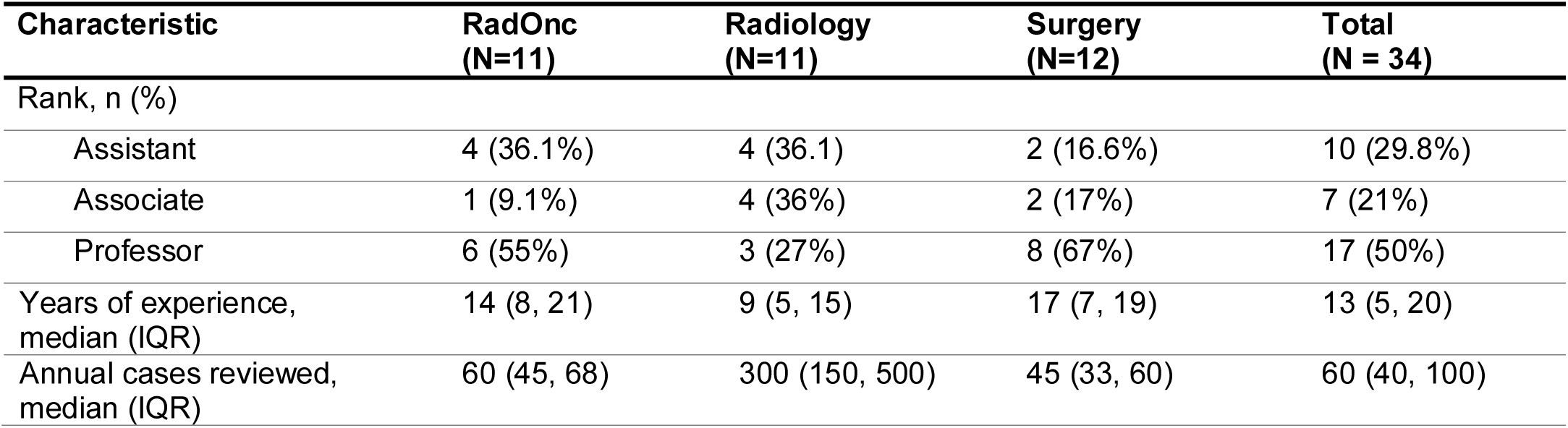
Clinician annotator demographic characteristics of the 34 physicians recruited for this study.

| Characteristic | RadOnc<br>(N=11) | Radiology<br>(N=11) | Surgery<br>(N=12) | Total<br>(N = 34) |
| --- | --- | --- | --- | --- |
| Rank, n (%) |  |  |  |  |
| Assistant | 4 (36.1%) | 4 (36.1) | 2 (16.6%) | 10 (29.8%) |
| Associate | 1 (9.1%) | 4 (36%) | 2 (17%) | 7 (21%) |
| Professor | 6 (55%) | 3 (27%) | 8 (67%) | 17 (50%) |
| Years of experience,<br>median (IQR) | 14 (8, 21) | 9 (5, 15) | 17 (7, 19) | 13 (5, 20) |
| Annual cases reviewed,<br>median (IQR) | 60 (45, 68) | 300 (150, 500) | 45 (33, 60) | 60 (40, 100) |

### Patient characteristics

Twenty-four patient cases with a pathologically confirmed diagnosis of HPV+ OPC were included in this analysis. All patients underwent lymph node dissection confirming pENE presence in 17 patients and absence in the remaining 7 patients. Patient demographics are shown in **Table 2**

**Table 2.**
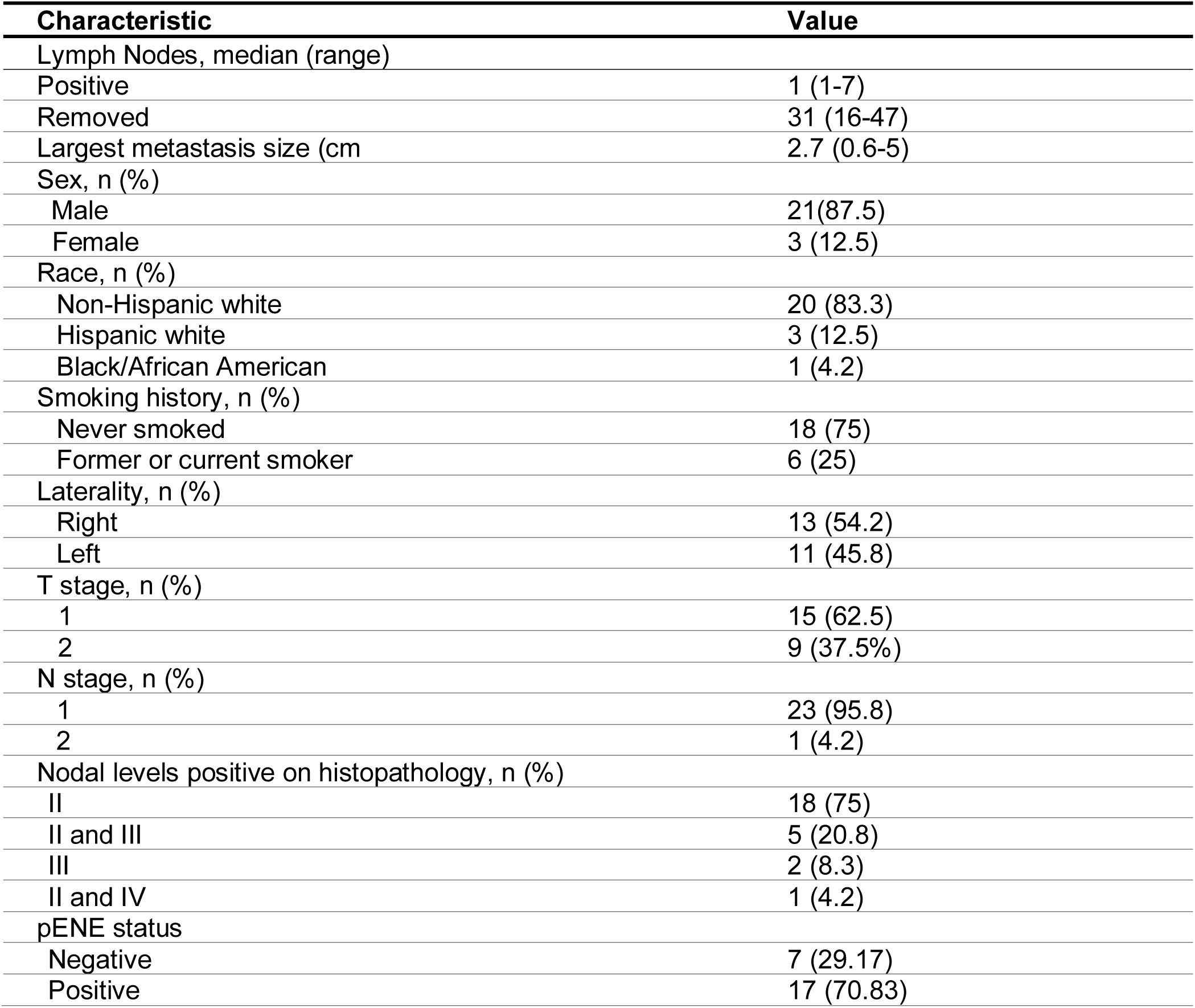
Patient demographic characteristics for the 24 OPC patients used in this study.

| Characteristic | Value |
| --- | --- |
| Lymph Nodes, median (range) |  |
| Positive | 1 (1-7) |
| Removed | 31 (16-47) |
| Largest metastasis size (cm) | 2.7 (0.6-5) |
| Sex, n (%) |  |
| Male | 21(87.5) |
| Female | 3 (12.5) |
| Race, n (%) |  |
| Non-Hispanic white | 20 (83.3) |
| Hispanic white | 3 (12.5) |
| Black/African American | 1 (4.2) |
| Smoking history, n (%) |  |
| Never smoked | 18 (75) |
| Former or current smoker | 6 (25) |
| Laterality, n (%) |  |
| Right | 13 (54.2) |
| Left | 11 (45.8) |
| T stage, n (%) |  |
| 1 | 15 (62.5) |
| 2 | 9 (37.5%) |
| N stage, n (%) |  |
| 1 | 23 (95.8) |
| 2 | 1 (4.2) |
| Nodal levels positive on histopathology, n (%) |  |
| II | 18 (75) |
| II and III | 5 (20.8) |
| III | 2 (8.3) |
| II and IV | 1 (4.2) |
| pENE status |  |
| Negative | 7 (29.17) |
| Positive | 17 (70.83) |

### Image acquisition and processing

De-identified pre-operative contrast-enhanced CT images were retrospectively acquired from the institutional picture archiving system in Digital Imaging and Communications in Medicine (DICOM) format. Patients underwent imaging following the standard institutional diagnostic head and neck CT imaging protocol using the following CT scanners: GE Discovery (n=16); GE Revolution (n=4); GE LightSpeed VCT (n= 3); and Siemens SOMATOM Edge Plus (n=1), with intravenous contrast administration. The kilovoltage peak was set at 120 kV for all patients with a median in-plane resolution of 0.49 mm (range: 0.49-0.53 mm), a slice thickness of 1.25 mm (range: 1.25-1.5 mm), an exposure time of 1000 ms (range: 1000-1825 ms), and X-ray tube current of 260 mA (159-409 mA).

CT images were converted to Neuroimaging Informatics Technology Initiative (NIfTI) format using the the DICOMRTTool v.3.2.0 Python package ^18^. All images were cropped to the cephalad border of the sternum and inferior border of the hard palate to exclude irrelevant anatomical regions. For intraobserver variability assessment, images from a random subset of 6 patients (4 with pENE, 2 without) were duplicated and randomly shuffled, resulting in a total of 30 cases: 21 with pENE and 9 without.

### Survey Instrument

Anonymized NIfTI images for the 30 cases were independently reviewed by observers using 3D Slicer ^19^ image-viewer accessed remotely via telemedicine software with remote control function enabled for image manipulation, scrolling and window-level setting (Supplementary **figure 1**). The observers answered a nine-question survey indicating presence or absence of seven iENE features: indistinct capsular contour, irregular lymph node margin, thick-walled enhancing nodal margin, perinodal fat stranding, perinodal fat plane or gross invasion, nodal necrosis, and nodal matting^20^. Additionally, observers predicted presence or absence of pENE and estimated their prediction confidence on a scale of 0-100% (**Appendix A**). Observers were blinded as to the results, as well as repeated images.

### Discriminative Performance Evaluation

Sample size justification was performed using the non-parametric method described by Pepe ^21^ with 30 planned observers of 24 independent cases, to detect an expected AUC of 0.70 with 1-β=0.8, and α=0.05. Discriminative performance was subsequently assessed using accuracy, area under the receiver operating characteristic curve (AUC), sensitivity, and specificity ^22,23^. Observer predictions were used to calculate accuracy, sensitivity, and specificity, while observer confidence scores were used to determine the AUC. All metrics are scaled from 0 to 1, with higher values indicating superior performance. Calibration of observer predictions was assessed using Brier score, also ranging from 0 to 1, with lower values indicating better calibration^24^. Performance was reported as medians with interquartile ranges (IQR). Mann-Whitney U tests were used to compare performance metrics between clinical specialties, and the Hanley and McNeil method was used to compare AUCs against the null (0.50) ^23^. All performance metrics were calculated in Python v.3.8.8 using the scikit-learn v.1.0.2 package ^25^; Mann-Whitney U tests were calculated using the *statannotations v.0.4.4* package^26^. The 95% confidence intervals were computed using a fast implementation of DeLong’s method via *confidenceinterval* package ^27,28^. *p* values less than or equal to 0.05 were considered significant.

### Radiographic Criteria Analysis

Sensitivity and specificity of the reported radiographic criteria for the correct identification of pENE were calculated across all observers and for each specialty. Logistic regression was performed using R version 4.2.2, to identify the significant radiographic features predictive of the true pENE status.

### Performance Variability Estimation

Inter-observer agreement for radiographic features among specialties was assessed by Fleiss’ Kappa using the irr v.0.84.1 package in R ^29,30^. Kappa values were interpreted following levels of agreement by Landis and Koch ^31^. To measure the reliability of the radiographic discriminative capacity of pENE by physicians, the intraclass correlation coefficient (ICC) was calculated using the pingouin v.0.5.3 package in Python. The standard error of measurement (SEm) was calculated using the duplicated cases to evaluate the intra-observer variability in pENE status assessment using the SEofM v.0.1.0 package in R was used to calculated the SEm ^32^.

## RESULTS

### Discriminative Performance

Median (IQR; 95%CI) performance aggregated across specialties demonstrated the following metrics: accuracy at 0.57 (0.10; 0.39 to 0.73), AUC at 0.64 (0.13; 0.44 to 0.83), Brier score at 0.28 (0.08; 0.44 to 0.83), sensitivity at 0.53 (0.27; 0.32 to 0.72), and specificity at 0.61 (0.33; 0.31-0.84).

Performance metrics aggregated by clinician specialty are shown in **Figure 1**. Surgeons had the highest median scores for accuracy (0.57), Brier score (0.26), and sensitivity (0.69). Radiation oncologists had the highest median scores for AUC (0.65), and specificity (0.89). There were significant differences between radiologists and surgeons for Brier score (0.33 vs. 0.26), radiation oncologists and surgeons for sensitivity (0.48 vs. 0.69), and radiation oncologists and radiologists/surgeons for specificity (0.89 vs. 0.56). The discriminative performance for the three specialties was not significantly different from random chance (**Figure 2**).

**Figure 1.**
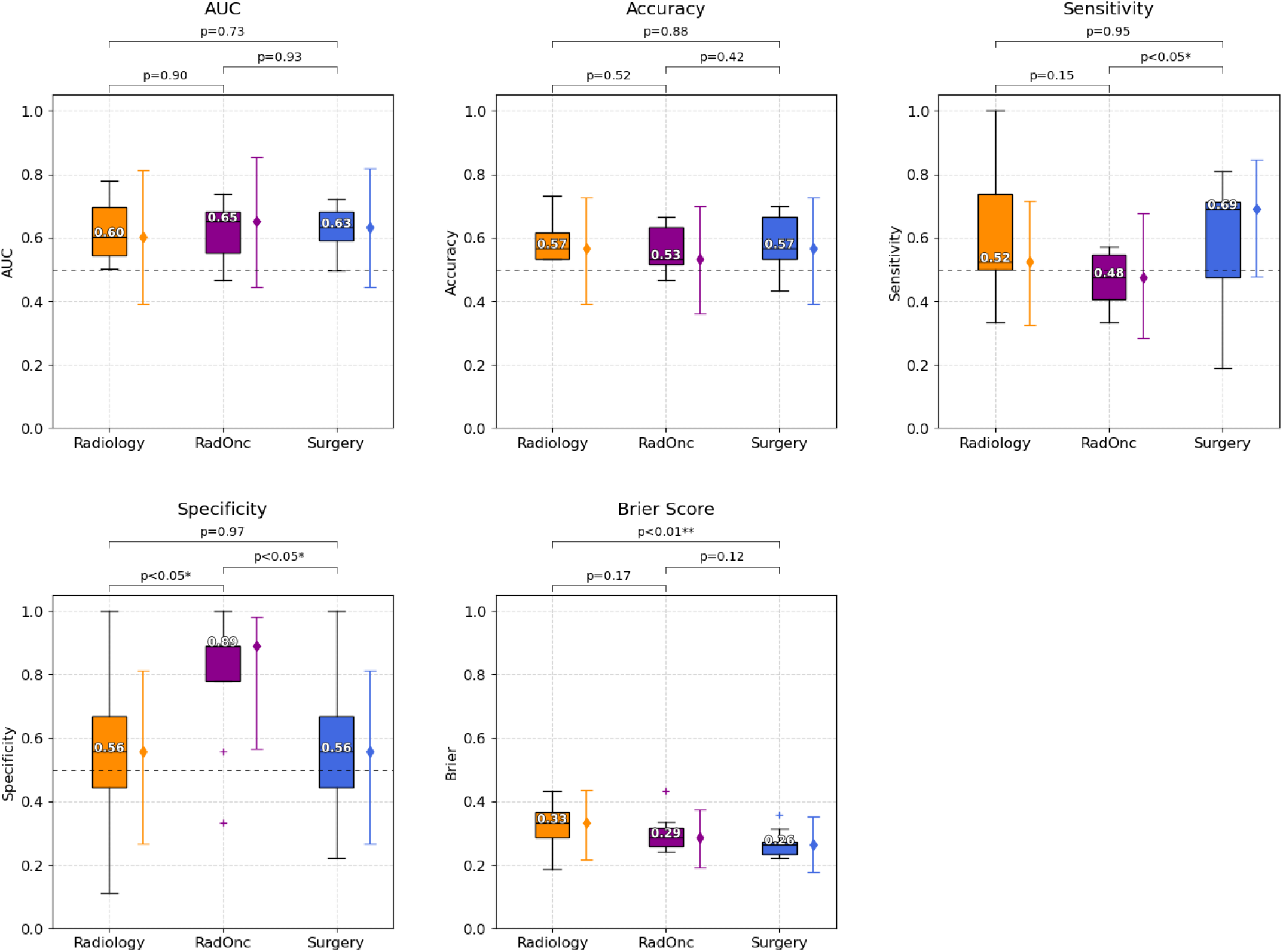
Comparisons of specialty-specific performance in detecting extranodal extension. Accuracy, sensitivity, and specificity, area under the receiver operating characteristic curve (AUC), and Brier scores are shown separately for radiologists (Radiology, orange color), radiation oncologists (RadOnc, purple color), and surgeons (Surgery, blue color). Higher values are deemed superior for all metrics except Brier score (where lower scores indicate better performance). Box plots represent the median (horizontal line within the box), and interquartile ranges (25th and 75th percentiles), with whiskers extending to the most extreme data points within 1.5 times the interquartile range from the box. The ‘+’ markers denote observations outside the range of adjacent values. Solid colored error bars represent the median (diamond marker) with 95% confidence intervals for each specialty. The horizontal dashed line at 0.5 on the AUC, accuracy, sensitivity, and specificity plots, is shown as reference line for threshold of no discrimination.

**Figure 2.**
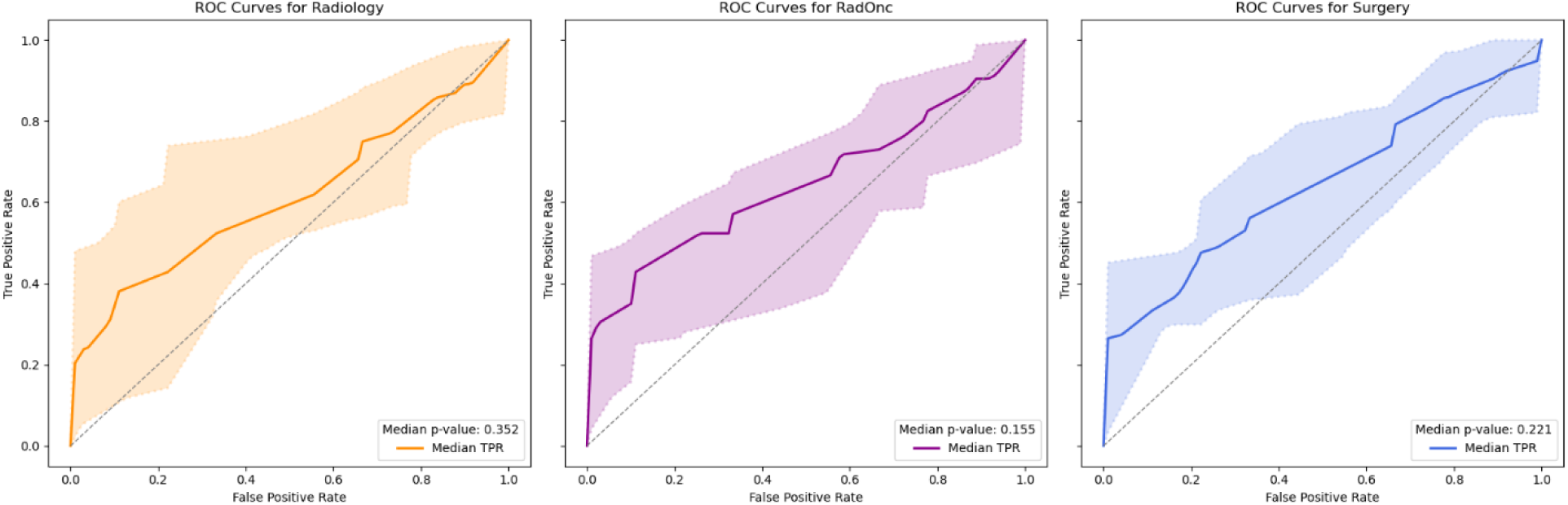
ROC Curves for Radiology, Radiation Oncology, and Surgery. The median p-values for the area under the curve (AUC) using the Hanley-McNeil method are shown. The shaded areas represent the 95% confidence intervals.

### Radiographic Criteria Analysis

Indistinct capsular contour (OR 1.71, p=0.01) and nodal matting (OR 1.5, p=0.02) emerged as significant predictors of pENE status among all physicians, with sensitivities of 82.8% and 62.6%, and specificities of 84.3% and 81.2%, respectively. (**Table 3**). Nodal necrosis was a strong negative predictor of pENE (OR 0.43, p<0.01). Assessment confidence was consistently associated with high odds ratios across specialties, especially for radiation oncologists (OR 3.77, p<0.01) and surgeons (OR 3.24, p<0.01).

**Table 3.** Logistic regression of correct ENE status prediction using radiographic features and assessment confidence. Significant p values: * = p ≤ 0.05, ** = p ≤ 0.01, *** = p ≤ 0.005.

|  | Coefficient | Std. Error | Odds Ratio | 95% CI |  | P-value | Sensitivity | Specificity |
| --- | --- | --- | --- | --- | --- | --- | --- | --- |
| **All Physicians** |  |  |  |  |  |  |  |  |
| Indistinct Capsular Contour | 0.54 | 0.21 | 1.71 | [1.14, 2.57] |  | 0.01 ** | 82.8% | 84.3% |
| Irregular Lymph Node Margin | 0.04 | 0.20 | 1.04 | [0.7, 1.55] |  | 0.83 | 87.1% | 74.6% |
| Thick-Walled Enhancing Nodal Margin | -0.33 | 0.18 | 0.72 | [0.51, 1.01] |  | 0.06 | 80.8% | 68.5% |
| Perinodal Fat Stranding | 0.09 | 0.17 | 1.1 | [0.78, 1.54] |  | 0.59 | 61.1% | 78.2% |
| Perinodal Fat Plane or Gross Invasion | 0.34 | 0.20 | 1.4 | [0.94, 2.08] |  | 0.09 | 62.9% | 92.9% |
| Nodal Necrosis | -0.84 | 0.19 | 0.43 | [0.3, 0.63] |  | <0.01 *** | 92.9% | 59.9% |
| Nodal Matting | 0.40 | 0.17 | 1.5 | [1.06, 2.1] |  | 0.02 * | 62.6% | 81.2% |
| Assessment Confidence | 1.00 | 0.15 | 2.73 | [2.04, 3.66] |  | <0.01 *** | - | - |
| **Radiologists** |  |  |  |  |  |  |  |  |
| Indistinct Capsular Contour | 1.01 | 0.37 | 2.74 | [1.33, 5.75] |  | 0.01 * | 79.7% | 89.3% |
| Irregular Lymph Node Margin | -0.04 | 0.37 | 0.96 | [0.46, 1.96] |  | 0.92 | 78.3% | 92.9% |
| Thick-Walled Enhancing Nodal Margin | 0.02 | 0.32 | 1.02 | [0.54, 1.89] |  | 0.96 | 82.6% | 76.8% |
| Perinodal Fat Stranding | 0.20 | 0.30 | 1.22 | [0.66, 2.20] |  | 0.52 | 74.6% | 64.3% |
| Perinodal Fat Plane or Gross Invasion | 0.81 | 0.38 | 2.24 | [1.07, 4.78] |  | 0.03 * | 60.1% | 96.4% |
| Nodal Necrosis | -0.59 | 0.38 | 0.55 | [0.26, 1.16] |  | 0.12 | 98.6% | 51.8% |
| Nodal Matting | 0.00 | 0.31 | 1.00 | [0.54, 1.84] |  | 0.99 | 56.5% | 80.4% |
| Assessment Confidence | 0.54 | 0.26 | 1.71 | [1.03, 2.86] |  | 0.04 * | - | - |
| **Radiation Oncologists** |  |  |  |  |  |  |  |  |
| Indistinct Capsular Contour | 0.34 | 0.38 | 1.41 | [0.66, 2.96] |  | 0.37 | 80.6% | 84.8% |
| Irregular Lymph Node Margin | 0.14 | 0.35 | 1.15 | [0.57, 2.31] |  | 0.69 | 87.0% | 72.2% |
| Thick-Walled Enhancing Nodal Margin | -0.43 | 0.31 | 0.65 | [0.35, 1.18] |  | 0.16 | 78.7% | 63.3% |
| Perinodal Fat Stranding | 0.18 | 0.36 | 1.20 | [0.58, 2.43] |  | 0.62 | 61.1% | 86.1% |
| Perinodal Fat Plane or Gross Invasion | -0.02 | 0.45 | 0.98 | [0.40, 2.34] |  | 0.97 | 69.4% | 96.2% |
| Nodal Necrosis | -0.68 | 0.32 | 0.51 | [0.27, 0.95] |  | 0.04 * | 88.0% | 59.5% |
| Nodal Matting | 0.48 | 0.35 | 1.62 | [0.82, 3.19] |  | 0.17 | 63.9% | 84.8% |
| Assessment Confidence | 1.33 | 0.33 | 3.77 | [2.02, 7.38] |  | <0.01 *** | - | - |
| **Surgeons** |  |  |  |  |  |  |  |  |
| Indistinct Capsular Contour | 0.28 | 0.36 | 1.32 | [0.65, 2.67] |  | 0.44 | 87.3% | 79.0% |
| Irregular Lymph Node Margin | 0.42 | 0.38 | 1.52 | [0.71, 3.24] |  | 0.28 | 95.3% | 61.3% |
| Thick-Walled Enhancing Nodal Margin | -0.56 | 0.30 | 0.57 | [0.31, 1.03] |  | 0.06 | 80.7% | 67.7% |
| Perinodal Fat Stranding | -0.06 | 0.29 | 0.94 | [0.53, 1.66] |  | 0.83 | 48.7% | 80.6% |
| Perinodal Fat Plane or Gross Invasion | 0.29 | 0.31 | 1.34 | [0.73, 2.43] |  | 0.34 | 60.7% | 85.5% |
| Nodal Necrosis | -1.40 | 0.36 | 0.25 | [0.12, 0.49] |  | <0.01 *** | 91.3% | 67.7% |
| Nodal Matting | 0.67 | 0.28 | 1.96 | [1.13, 3.41] |  | 0.02 * | 67.3% | 77.4% |
| Assessment Confidence | 1.18 | 0.26 | 3.24 | [1.98, 5.42] |  | <0.01 *** | - | - |

### Performance Variability

Inter-observer agreement for radiographic features was generally weak (supplementary figure 2). Among radiologists, agreement was weak (0.4 < Kappa < 0.6) when assessing thick-walled enhancing nodal margin, nodal necrosis, perinodal fat stranding, and indistinct capsular contour, and minimal (0.2 < Kappa < 0.4) for irregular lymph node margin, perinodal fat plane or gross invasion, and nodal matting. Radiation oncologists also showed weak agreement in evaluating nodal matting and thick-walled enhancing nodal margin, and minimal agreement in all other features. Surgeons had weak agreement only in assessing thick-walled enhancing nodal margin, no agreement (0 < Kappa < 0.2) in perinodal fat stranding and perinodal fat plane or gross invasion, and minimal agreement in all other radiographic features. The SEm demonstrated that interobserver variability was generally higher than intraobserver variability, with surgeons showing the highest variability (supplementary figure 3). The intraclass correlation coefficient (ICC) for all physicians was 0.36 (95% CI = [0.26, 51], p < 0.01).

## DISCUSSION

In this study, we queried a large number of clinicians across three key specialties involved in the management of HPV+ OPC patients to determine differences in the detection of iENE and prediction of pENE status. Broadly, we determine that though differences do exist between specialists, the overall ability of clinicians to correctly predict pENE using CT imaging was uniformly poor. To our knowledge, this is the largest individual prospective blinded human performance study to investigate radiological interpretation for ENE in HPV+ HNSCC across multiple specialties.

Our study aligns with previous research demonstrating poor discriminative performance and high variability among radiologists in identifying pENE using pre-operative CT imaging. For example, a recent meta-analysis reported pooled sensitivity, specificity, and AUC values of 0.77, 0.60, and 0.72, respectively, for CT-based identification of ENE in OPC ^33^. While our aggregated values are notably lower for sensitivity (though still within the 95% confidence interval), our specificity and AUC are similar. Interestingly, our study found that radiation oncologists had significantly higher specificity than the other specialties, suggesting they are more accurate in identifying true negative cases who are less likely to benefit from CRT. On the other hand, surgeons demonstrated the most reliable Brier scores across all specialties with significantly lower values, i.e. better calibration, than radiologists. This is likely due to more conservative estimates of confidence, i.e., avoiding overconfidence in uncertain cases and vice versa. These findings highlight significant implications for clinical practice and suggest that the variable performance and prediction of pENE may be related to specialty-specific heuristics. For example, the higher sensitivity but lower specificity exhibited by surgeons and radiologists indicates a tendency to err on the side of caution, which results in forgoing surgery in ambiguous cases, likely to minimize toxicities associated with triple modality treatment. Conversely, radiation oncologists’ higher specificity suggests a more conservative approach with more selective recommendation of CRT.

In a large-scale meta-analysis for all HNSCC subtypes, it was found that central node necrosis showed high pooled sensitivity, while infiltration of adjacent planes showed high pooled specificity ^34^. These findings are echoed in our study as nodal necrosis was the most observed feature in aggregate for correctly determining ENE presence, while perinodal fat plane or gross invasion was the least commonly observed feature for correctly determining ENE absence. It should be noted that nodal necrosis was observed in almost all cases where ENE was correctly identified and in a sizable portion of cases where ENE was correctly ruled out, as could be expected for HPV+ OPC ^35^. For surgeons, rather than nodal necrosis, irregular lymph node margin was the most observed criterion for correct identification of ENE presence, which may be linked to their high sensitivity. Notably, on regression analysis, several radiographic criteria were significant contributors to the correct determination of ENE status. Moreover, there were some differences that emerged in significant criteria when stratifying the regression analysis by clinician specialty. However, irregular lymph node margin, thick-walled enhancing nodal margin, and perinodal fat stranding were among the criteria not deemed significant. This is not necessarily surprising given that these criteria have been less routinely reported in ENE studies ^12,33,34^.

Recent literature in HPV+ OPC ENE identification has suggested that CT radiographic criteria have poor reproducibility among expert observers ^12^, though there could be some improvements in reproducibility when using a high certainty threshold for ENE identification, consolidating operational definitions, and the sharing of experience among observers ^36^. We sought to determine if these findings were consistent when stratified by clinician specialty. Notably, Fleiss’ kappa was always less than 0.6, regardless of specialty or radiographic criteria, consistent with findings from Tran et al. ^12^. As expected, radiographic features that had higher agreement, both overall and within specialties, tended to have lower intraobserver and interobserver variability. Additionally, though there were features with relatively high agreement and low intra/interobserver variability, it is not clear if these features can be used to predict ENE as their presence may not be significantly associated with the correct prediction of ENE, as seen with thick-walled enhancing nodal margin ^34^.

Our study is not without limitations. First, we only investigated a single imaging modality for the identification of ENE status, namely CT. While recent evidence has suggested the incorporation of additional imaging modalities, such as magnetic resonance imaging (MRI) and semi-quantitative positron emission tomography (PET) parameters, could improve the discrimination of ENE in OPC ^33,37–39^, CT is among the most ubiquitous diagnostic imaging modalities available for OPC patients. Therefore, we have chosen to focus on CT as an exemplar imaging modality in this study. Secondly, due to not all patients having complete pathological ground truth information for ENE extent, we did not utilize this as a factor in our analysis. However, it is well known that depending on the ENE extent (i.e., > 2 mm), discriminant capacity often increases ^10^. Finally, while most patients in this dataset only had one positive lymph node, some patients with multiple positive nodes could have added unaccounted for ambiguity in clinician determination of ENE status. Additionally, while pathologic assessment of ENE was used as a gold standard for this study, the accuracy of this assessment method has been questioned in the literature ^40–42^.

The observed poor discriminative performance and limited agreement denoted herein have substantive implications for head and neck treatment selection in OPC. We have previously shown that the optimal policy for selecting initial therapy or radiation therapy for toxicity minimization for OPC is highly driven by the expected probability of ENE, and thus the need for TORS+ND patients to receive adjuvant radiotherapy ^4^. Our data herein show that humans, regardless of specialty, cannot routinely predict ENE, and therefore are largely incapable of accurately risk assessing for optimal side effect sparing via TORS+ND. This is coherent with multiple reported series whereby surgical patients dispositioned to TORS+ND to evade radiation-related sequelae in fact require adjuvant radiotherapy, or tri-modality (surgery/chemotherapy/radiation) owing to pathologically observed ENE, even when explicitly radiographically overt ENE cases are excluded ^5,15,43^. In sum, most patients dispositioned to surgery with the intent of evading radiotherapy with extant radiographic lymphadenopathy appear to be selected by an largely optimistic and inaccurate heuristic, rather than a reproducible assessment.

The inter-observer agreement reported in this study aligns with previous findings of moderate agreement levels of iENE ^33,44^. Our findings suggest that the observed variability in performance metrics across specialties may stem from inherent uncertainties in field-specific training and interpretative practices. It is still reasonable to assume potential value for the incorporation of lexicons and certainty levels to enhance inter-observer consistency of iENE as reported in a recent study^45^. Yet, whether this translates to improvements in real clinical settings remains to be determined.

Overall, our study reinforces the findings of previous investigations, which caution against relying solely on human interpretation of iENE from radiological imaging as a predictor of pENE. Given the difficulty of iENE/pENE detection for human observers regardless of clinical specialty, even when utilizing defined radiographic criteria, it is pertinent that solutions are put forth that could improve or automate this task. In recent years, machine learning approaches have been proposed as accurate and reproducible tools for determining ENE status from radiological images of HNSCC patients ^46–48^. We anticipate these methods to play an increasing role in the clinical utility of radiological determination of OPC ENE status in the future.

## CONCLUSIONS

In summary, prospectively assessing inter-/intra-rater/specialty human discriminant performance by querying 34 clinician annotators across 30 HPV+ OPC cases using a rigorous blinded studyshows that t radiologists, radiation oncologists, and surgeons have similarly poor discrimination of ENE status as determined through various evaluation metrics. Moreover, there was high variability between and within specialties. Put simply, human expert observers do not seem capable of reliably predicting pENE status, and therefore, effective allocation to surgical or non-surgical therapy is pre-empted by lack of effective prediction of ENE-directed adjuvant therapy. Future studies should incorporate the utilization of additional complementary imaging modalities (e.g., MRI and PET) and/or automated approaches (e.g., machine learning) that would improve discriminative performance and minimize variability of iENE identification.

## Data availability statement

In accordance with the *Final NIH Policy for Data Management and Sharing*, NOT-OD-21-013, anonymized tabular analytic and NIFTI data that support the findings of this study are openly available in an NIH-supported generalist scientific data repository (figshare) at http://doi.org/10.6084/m9.figshare.22177574 no later than the time of an associated peer—reviewed publication; while public data is embargoed pending peer review, the data is available upon request pre-peer-review through email to the corresponding author.

## Pre-print availability statement

In accordance with NIH Policy NOT-OD-17-050, *Reporting Preprints and Other Interim Research Products*, which specifies: “The NIH encourages investigators to use interim research products, such as preprints, to speed the dissemination and enhance the rigor of their work”, we have deposited a pre-peer review version of this manuscript on the medrxiv.org preprint server at https://doi.org/10.1101/2023.02.25.23286432.

## CRediT statement

In accordance with the Contributor Roles Taxonomy (CRediT, https://credit.niso.org/), the contributing authors have designated responsibilities and individual author attribution. The corresponding authors (ACM, CDF) assume responsibility for role assignment, and all contributors have been given the opportunity to review and confirm assigned roles: **Conceptualization:** OS, KAW, DIR, DS, ASRM and CDF**; Data curation:** OS; **Formal analysis:** OS, SK, KAW and CDF; **Funding acquisition:** KAW and CDF; **Investigation:** OS, SK, KAW, NT, RH, MAN, SS, AM, BHK, KK, JS, JJ, MA, MMC, GMC, EMD, ASG, RPG, JPG, GBG, JH, FH, NGT, JJ, DK, SDK, KN, SYL, ML, KOL, AL, CML, AM, ACM, JNM, JP, KBP, DIR, VS, DS, SJS, AGS, MW, ASRM, and CDF; **Methodology:** OS, SK, KAW and CDF; **Project administration:** OS and CDF; **Resources:** OS, SK, KAW, NT, RH, MAN, SS, AM, BHK, KK, JS, JJ, MA, MMC, GMC, EMD, ASG, RPG, JPG, GBG, JH, FH, NGT, JJ, DK, SDK, KN, SYL, ML, KOL, AL, CML, AM, ACM, JNM, JP, KBP, DIR, VS, DS, SJS, AGS, MW, ASRM, and CDF; Software: OS; **Supervision:** CDF; **Validation:** OS; **Visualization:** OS, SK; **Writing – original draft:** OS, SK, KAW and CDF**; Writing - review & editing:** OS, SK, KAW, NT, RH, MAN, SS, AM, BHK, KK, JS, JJ, MA, MMC, GMC, EMD, ASG, RPG, JPG, GBG, JH, FH, NGT, JJ, DK, SDK, KN, SYL, ML, KOL, AL, CML, AM, ACM, JNM, JP, KBP, DIR, VS, DS, SJS, AGS, MW, ASRM, and CDF.

## ICJME author statement

In accordance with International Committee of Medical Journal Editors (ICJME, https://www.icmje.org/) recommendations, all authors affirm qualification for authorship via the following criteria: *“Substantial contributions to the conception or design of the work; or the acquisition, analysis, or interpretation of data for the work; AND Drafting the work or reviewing it critically for important intellectual content; AND Final approval of the version to be published; AND Agreement to be accountable for all aspects of the work in ensuring that questions related to the accuracy or integrity of any part of the work are appropriately investigated and resolved.”*

## Data Availability

Raw data are available from authors upon reasonable request. Anonymized datasheets used for analysis will be made publicly available on Figshare (DOI = 10.6084/m9.figshare.22177574) after manuscript acceptance in peer reviewed journal.

https://doi.org/10.6084/m9.figshare.22177574

## Supplementary Material

**Supplementary figure 1.**
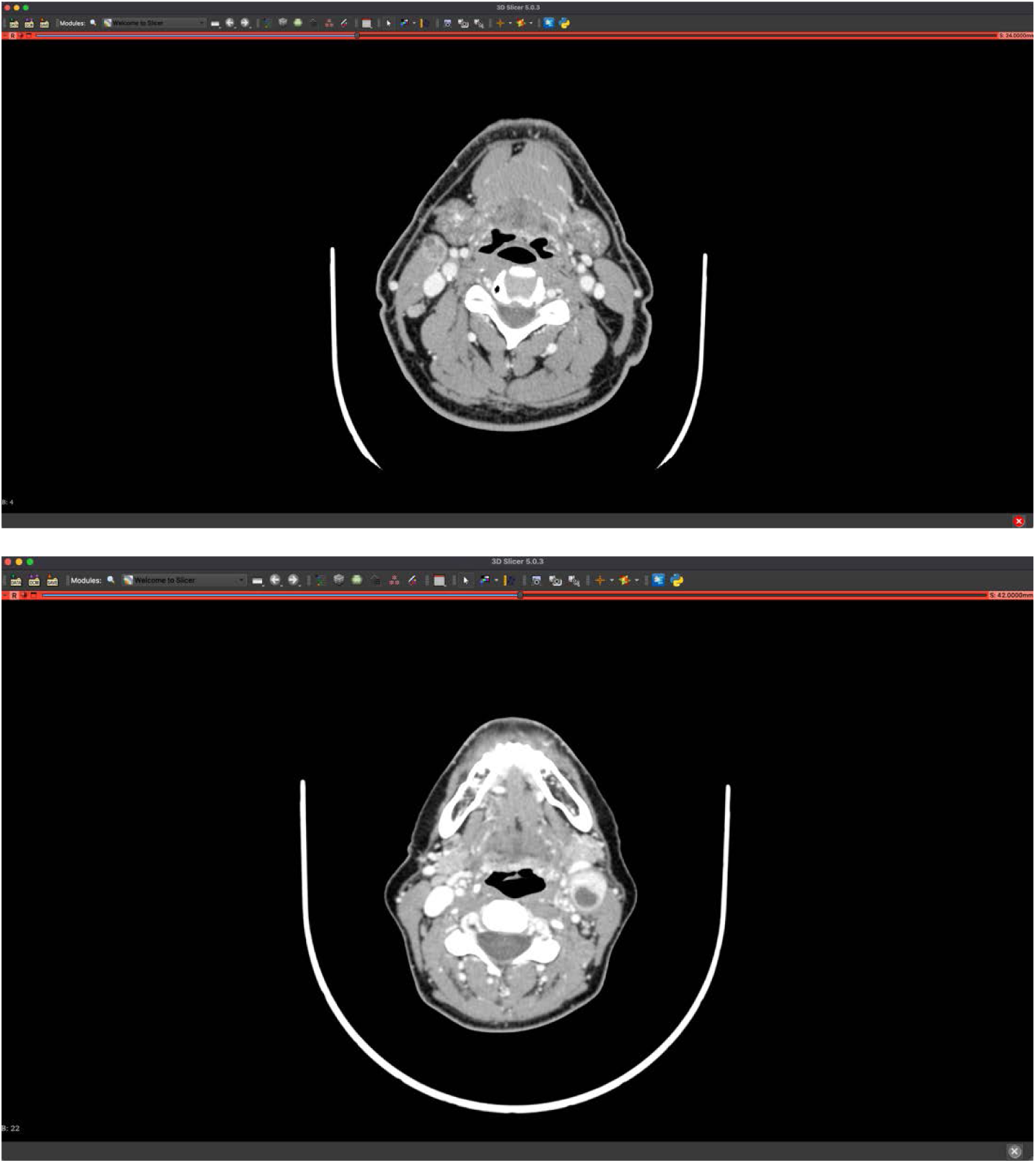
Example of CT scan in 3D Slicer with (top) and without (bottom) ENE presence as seen by observers. Observers could scroll through the scan remotely, change planes between axial, sagittal, or coronal, and change the window level and width.

**Supplementary figure 2.**
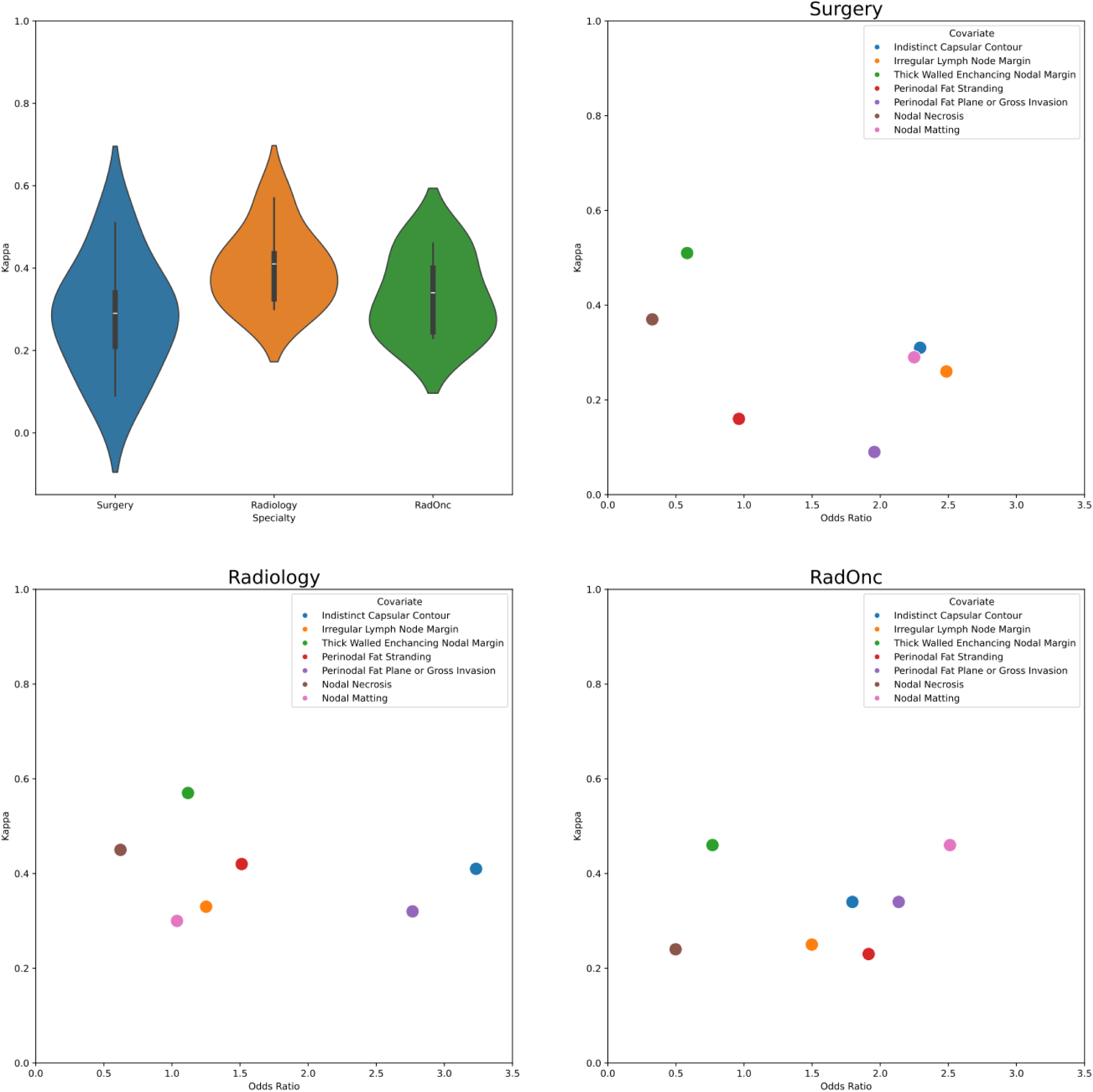
Fleiss’ Kappa for the seven radiographic features for each specialty. Higher values represent greater agreement in the evaluation of presence or absence for each feature. Subplots show agreement versus odds ratio in correctly determining ENE for each feature stratified by clinician specialty. The top right corner of the subplots represents features with high agreement and high predictive value.

**Supplementary figure 3.**
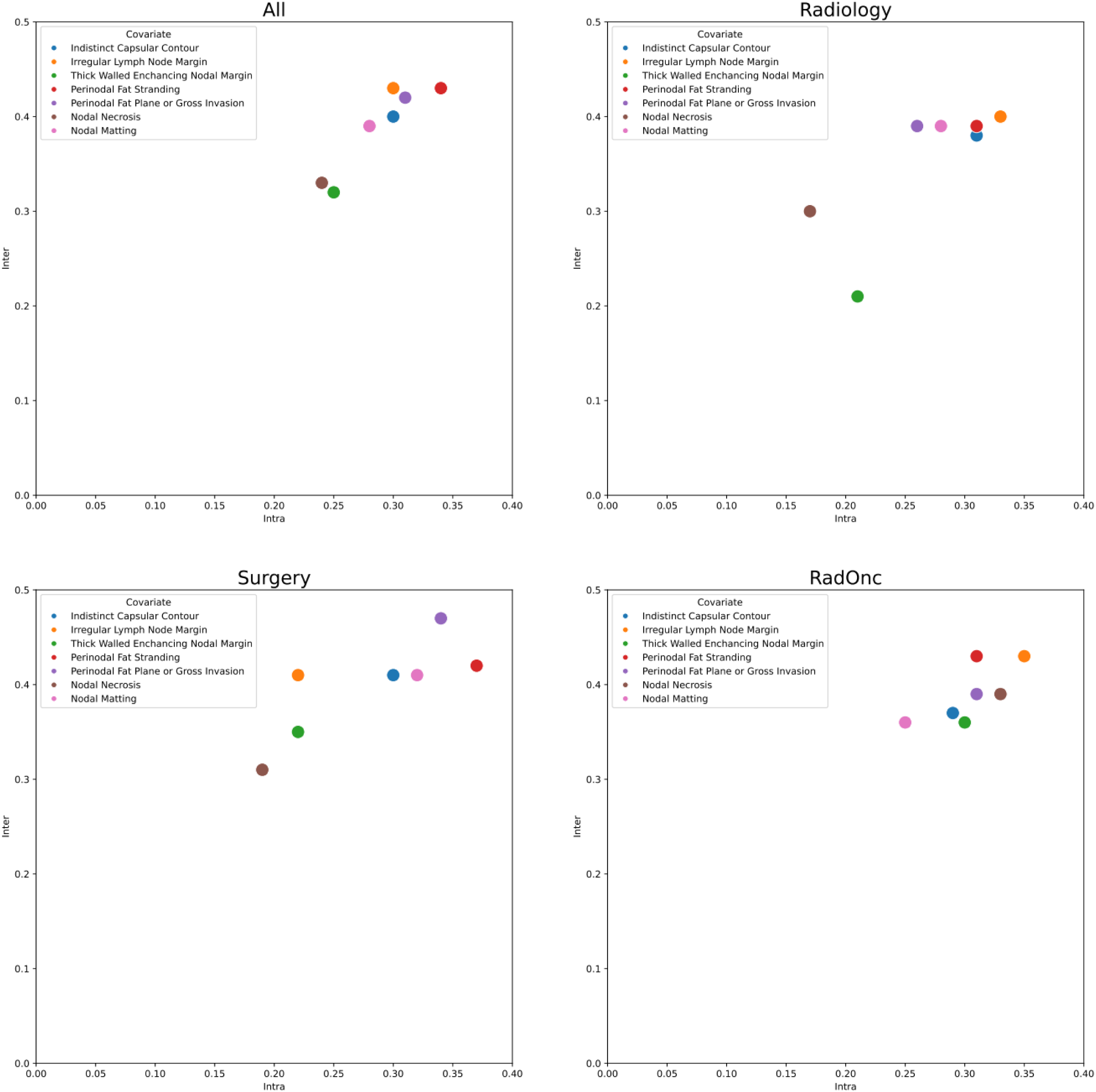
Interobserver vs. intraobserver variability plots as measured with the standard error of measurement. Each colored dot corresponds to a radiographic criterion. Results are presented for all observers and stratified by clinician specialty. Values in the bottom left corner represent features with low interobserver variability and low intraobserver variability, so would be preferred.

